# Who is wearing a mask? Gender-, age-, and location-related differences during the COVID-19 pandemic

**DOI:** 10.1101/2020.07.13.20152736

**Authors:** Michael H. Haischer, Rachel Beilfuss, Meggie Rose Hart, Lauren Opielinski, David Wrucke, Gretchen Zirgaitis, Toni D. Uhrich, Sandra K. Hunter

## Abstract

Masks are an effective tool in combatting the spread of COVID-19, but some people still resist wearing them and mask-wearing behavior has not been experimentally studied in the United States. To understand the demographics of mask wearers and resistors, and the impact of mandates on mask-wearing behavior, we observed shoppers (n = 9935) entering retail stores during periods of June, July, and August 2020. Approximately 41% of the June sample wore a mask. At that time, the odds of an individual wearing a mask increased significantly with age and was also 1.5x greater for females than males. Additionally, the odds of observing a mask on an urban or suburban shopper were ~4x that for rural areas. Mask mandates enacted in late July and August increased mask-wearing compliance to over 90% in all groups, but a small percentage of resistors remained. Thus, gender, age, and location factor into whether shoppers in the United States wear a mask or face covering voluntarily. Additionally, mask mandates are necessary to increase mask wearing among the public to a level required to mitigate the spread of COVID-19.

## Introduction

Wearing a mask in public is currently a controversial and politicized issue in the United States, even with case evidence from other countries that face coverings help to control the spread of coronavirus disease 2019 (COVID-19) [1]. As Taiwan, Hong Kong, South Korea, and other countries with almost universal masking have been some of the most successful at reducing daily case and death rates [2,3], the United States is trending in the other direction, setting national and state records for daily new cases of COVID-19 at the end of June 2020. Despite the controversy, COVID-19 is a virus that can be transmitted through respiratory droplets from infected individuals and even asymptomatic cases can be contagious [4]. Thus, without a vaccine in distribution, masks are one of the few control measures available for protection against the virus because they serve as a physical barrier between people [5,6]. Not only do masks protect the wearer from aerosolized droplets, but they also provide source control, stopping large droplets coming from a wearer before they become airborne. Study of the filtration efficiency of different fabrics has shown that even cotton weaves and blends can block viral transmission [7], so masks can be made virtually cost-free with household materials.

Though the evidence of the efficacy and cost-effectiveness of masks is clear, store policies and public mandates requiring masks in America have been met with protests [8] and, in rare cases, violence [9,10]. Public health research shows these measures may already have reduced cases in the United States by 450,000 through May 22^nd^ [11], but the messaging from top-level government officials has been inconsistent [12] and polling suggests that a sizeable portion of the general population are still going out in public without masks [13,14], though self-reported behavior is not always reliable. Women report wearing masks more often than men [14], but demographic differences in mask wearing have not been directly observed. Older adults are at higher risk for more severe cases of COVID-19 [15], so it is reasonable to expect that this demographic may wear masks more than younger individuals. Mask wearing by retail shoppers may also vary by location, in that groups that report greater resistance to mask wearing may be more concentrated outside of urban areas [16]. Specifically, rural areas tend to have higher concentrations of conservative-leaning voters [17] who are reported to be more resistant to wearing a mask [16]. Learning what demographic groups are wearing masks and how mask mandates impact behavior among these groups will help officials to better target public health messaging that promotes the practice among groups with lower usage. To facilitate greater understanding and reliable experimental data on whether gender, age, location, and the presence of mask mandates influence mask wearing in the United States, we conducted a direct observational study at retail stores in Wisconsin. Observations occurred between June and August 2020. From June 9^th^ to August 3^rd^, 2020, the 7-day average case rate in Wisconsin more than doubled (341 to 844 cases) and the percentage of positive cases had increased from 3.5% to 7.4% [18]. Thus, during this period, many major retail chain [19] and Wisconsin state [20] mask mandates were enacted, allowing for study of mask wearing before and after these regulations went into effect.

The primary aim of our study was to determine mask use by gender expression, estimated age, and location. We hypothesized that, without a mandate, mask use among females would be greater than males across all age groups and locations. Additionally, that mask use would be greater among older adults (>65 years old) than middle-age (30-65 years old) and young individuals (2-30 years old) and would be less in rural than in urban or suburban areas. A secondary aim of this project was to revisit stores on dates near the implementation of store and state mask mandates to characterize how mask-wearing behavior changes in response to these requirements.

## Materials and Methods

We addressed our primary aim by visiting 36 different retail locations across five different counties in southeastern Wisconsin. Retail stores selected for observation were based largely on geographical spread across Milwaukee and the surrounding areas. In the interest of exploring mask wearing among urban, suburban, and rural stores, we designated a center point of the city (United States Postal Service main office; 345 W St. Paul Ave, Milwaukee, WI, USA) and recorded the linear distance of each store to the city center. Based on distance, stores were then placed into urban (<6.1 km from city center; *n* = 15, 3.3 ± 1.8 km to city center), suburban (11.5-32.1 km; *n* = 12, 20.5 ± 7.2 km), and rural (>36.9 km; *n* = 9, 55.8 ± 21.4 km) store groups. Visits to stores (*n* = 36) occurred at various times between 9am and 8pm (June 3^rd^-9^th^, 2020) and typically lasted less than an hour, with observers recording data with writing utensil and paper or mobile device. To address our secondary aim, stores were revisited and data was collected in the same manner on dates shortly before store or state mandates were implemented (*n* = 8; July 24^th^-31^st^, 2020), when there was a store mandate but not a state mandate (*n* = 20; July 22^nd^-July 31^st^, 2020), and after the Wisconsin state mandate began (*n* = 10; August 1^st^-3^rd^, 2020). Note that the overlap in the secondary visit dates is related to corporate differences in the dates mask mandates began at various stores. Stores consisted of grocery and other large big-box retail stores and shoppers were not aware they were being observed. Children under the estimated age of two were not recorded. For all observations that were collected, summary sheets were crosschecked by other observers. All procedures involved public observation or accessed public information and did not require review by an Institutional Review Board.

To determine the impact of gender, age, location, and their interactions on mask wearing during the initial visit period (June 3^rd^-9^th^, 2020), multiple logistic regression analysis was performed. Sixty individuals who were wearing their mask or face covering improperly (not over nose and mouth) were recorded but excluded from this analysis as they could not be grouped into a dichotomous outcome (mask/no mask). 5517 observations remained for this analysis, and gender expression, age, and location independent variables were dummy coded and entered into a backward elimination procedure with their associated interactions (gender-age, gender-location, age-location, and gender-age-location). Limit for variable removal and test classification cutoff were set at 0.025 and 0.5, respectively. Adjusted odds ratios (aOR) are expressed with respect to reference groups (gender: male, age: young, location: rural) with 95% confidence intervals and significance was determined at *p* < 0.05.

Percentage data including incorrect mask wearers from the initial visit (June 3^rd^-9^th^, 2020) was used to compare to later data (July 24^th^-Aug 3^rd^, 2020) and evaluate the changes in mask-wearing behavior in response to mask mandates. All analyses were performed with either Microsoft Excel (Microsoft, Redmond, WA, USA) or IBM Statistical Package for Social Sciences version 26 (IBM, Armonk, NY, USA).

## Results and Discussion

Over the course of 74 visits, 9935 individuals were observed entering retail stores. Of the 5517 individuals we observed during the first round of data collection that could be grouped into a dichotomous outcome (mask/no mask) (Fig 1), 41.5% were wearing a mask or face covering with the general trend across gender, age, and location largely aligning with our original hypotheses (Table 1). Females wore masks 7.6% more than males (Fig 2A). Additionally, masks were seen at a higher percentage in older than middle-age (+16.1%) and young (+19.8%) individuals (Fig 2B). Urban and suburban mask wearing was similar but was much lower at rural stores (Suburban: −28.7%; Urban: −26.4%; 55.81 ± 21.37 km) (Fig 2C).

**Table 1.** Observed odds 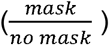 of wearing a mask by age and location during initial data collection period from June 3^rd^-9^th^, 2020*

|  | *Young* |  | *Middle Age* |  | *Older* |  |
| --- | --- | --- | --- | --- | --- | --- |
|  | Female | Male | Female | Male | Female | Male |
| *Urban* | 0.879 | 0.639 | 1.022 | 0.863 | 2.115 | 1.049 |
| *Suburban* | 0.886 | 0.567 | 1.257 | 0.763 | 2.113 | 1.468 |
| *Rural* | 0.076 | 0.117 | 0.287 | 0.172 | 1.094 | 0.600 |

\* Odds > 1 show odds of wearing a mask are greater than not wearing a mask

**Fig 1.**
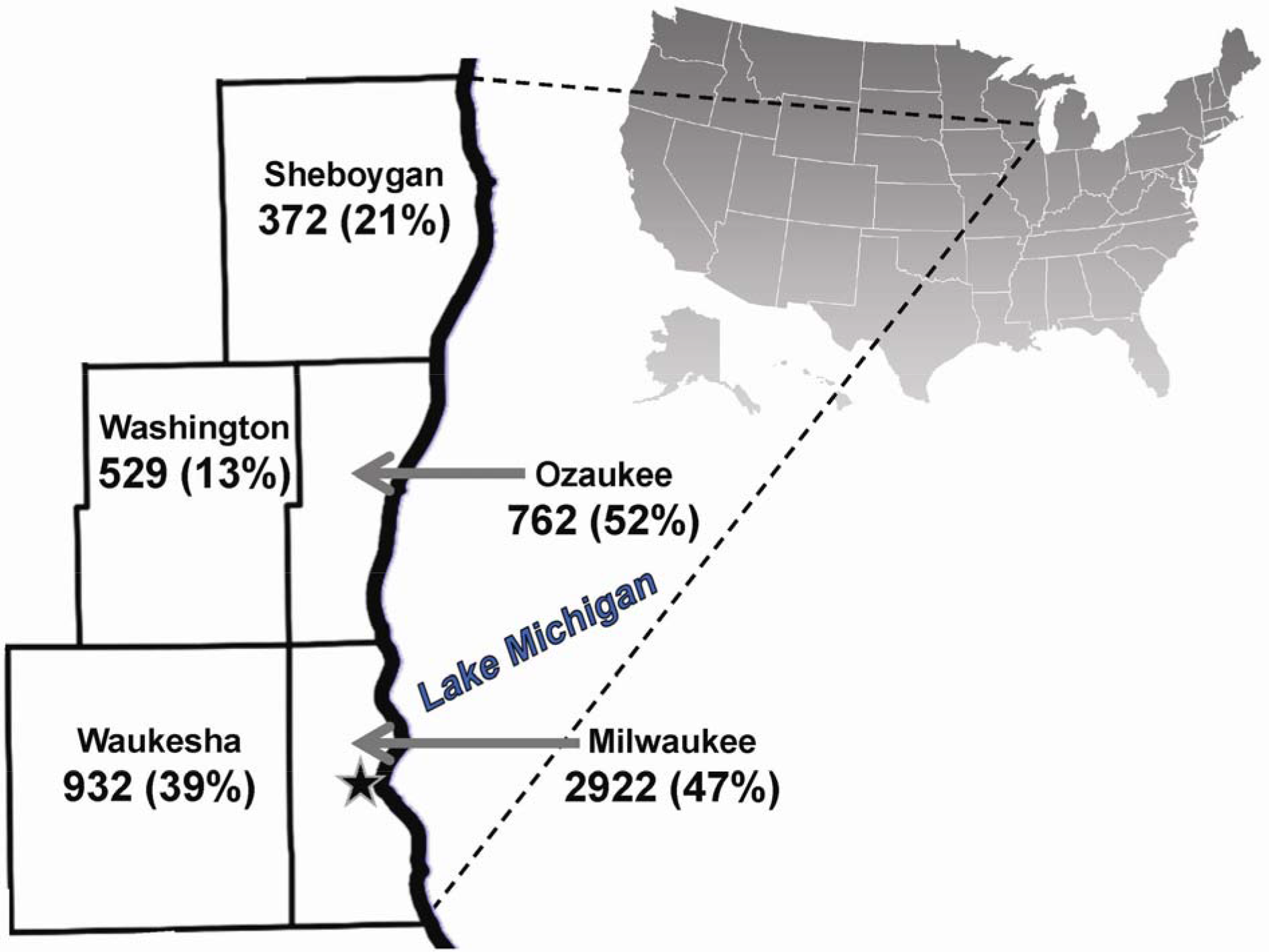
Mask-wearing percentages by Wisconsin county. Data shown indicates the number of observations collected and percentage of people wearing a mask (vs. no mask) in each county where retail stores were visited during the initial data collection period from June 3^rd^ through June 9^th^, 2020. The star symbol in Milwaukee county represents the city center location used for identifying urban, suburban, and rural stores.

**Fig 2.**
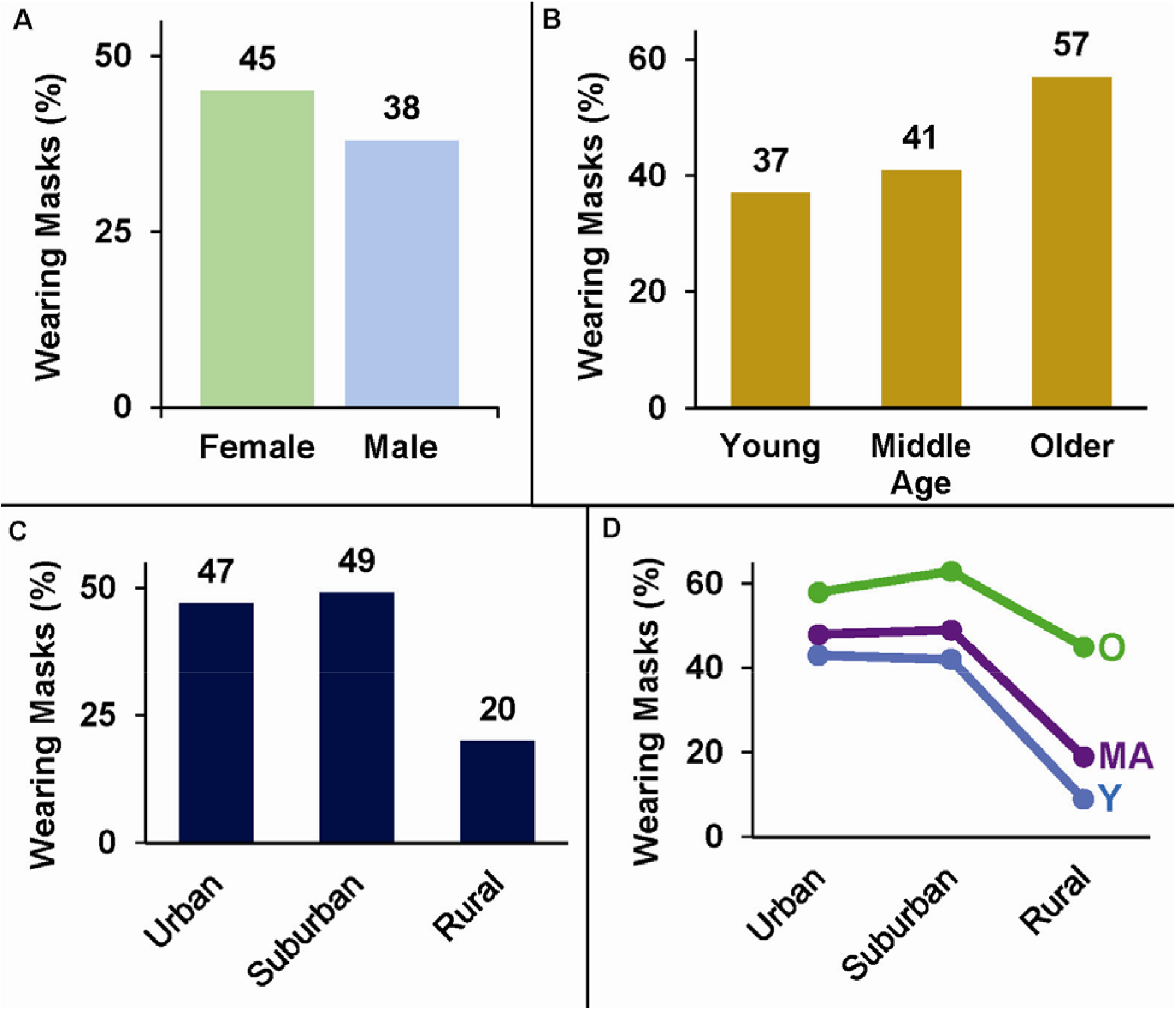
Mask-wearing percentages across gender, age, and location. Data shown indicates percentage of individuals wearing a mask (vs. no mask) during the initial data collection period from June 3^rd^ through June 9^th^, 2020. **A**. Females wear masks more than males. **B**. Older adults wear masks more than other individuals. **C**. Mask-wearing habits are similar in urban and suburban areas, but usage drops off considerably at rural stores. **D**. Mask wearing plotted by location highlights how the mask-wearing behavior of older adults (O) is less impacted by shopping in a rural area than the behavior of young (Y) and middle-age (MA) individuals.

Regression analysis revealed that gender, age, and location all significantly impact the odds of an individual being observed to wear a mask (p < 0.001; Fig 3). Results indicate the odds of a female wearing a mask are significantly greater than males (aOR = 1.470, 95% CI = 1.313-1.646). Odds are also greater for middle age (aOR = 1.597, 95% CI = 1.359-1.877) and older adults (aOR = 3.434, 95% CI = 2.811-4.195) than younger individuals. The odds of observing someone in urban (aOR = 3.847, 95% CI = 3.157-4.689) or suburban (aOR = 4.124, 95% CI = 3.418-4.975) areas wearing a mask is also much higher than in rural locations, reflecting the much lower prevalence of masks seen at rural stores. The significant age-location interaction effect (p < 0.001) is largely driven by differential changes in mask-wearing behavior between older adults and the other age groups (Fig 2D).

**Fig 3.**
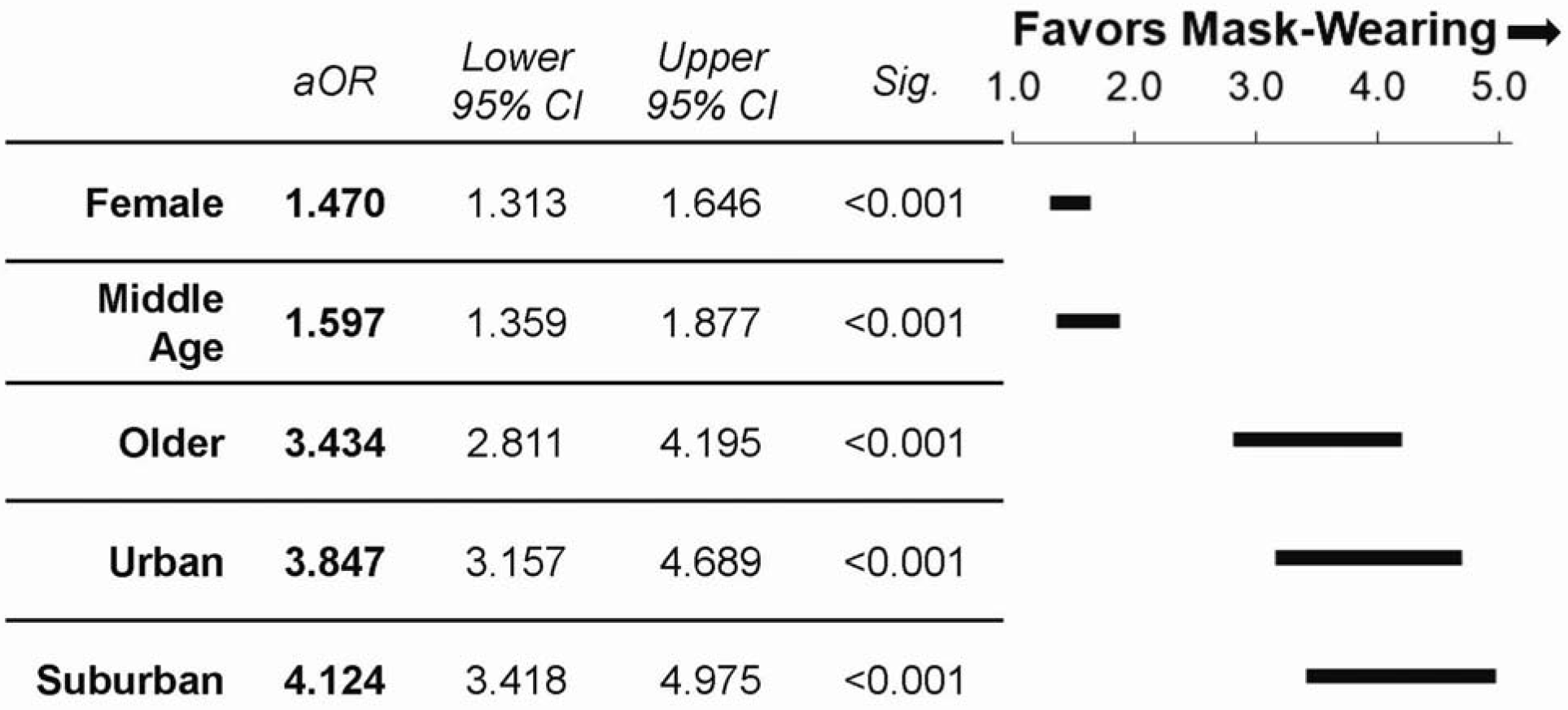
Odds of wearing a mask. Adjusted odds ratios (aOR) and 95% confidence interval plots of mask usage for gender expression, age, and location during the initial data collection period from June 3^rd^-9^th^, 2020. Odds ratios are expressed in relation to reference groups (gender: male; age: young; location: rural).

Currently, no peer reviewed and direct observational studies of mask wearing are available for comparison to our study findings. Preprint study akin to ours also reflects alarming and similarly low percentages of mask wearing in Wisconsin [21]. Additionally, our data can be discussed in relation to national polls and surveys that addressed mask wearing at about the same time. A May-to-June survey of adults in Wisconsin and four other surrounding states indicated 45.1% always wear a mask when they leave home [13], and data in our study lends credibility to this number. Notably, other regions of the United States surveyed reported different habits (33.5-64% mask usage) so direct observational studies in other parts of the country would be beneficial to confirm national variability when mask mandates are not in place. Another poll conducted in mid-April indicated that only 36% of adults always wear a mask or face covering when outside the home [14], so perhaps usage nation-wide has gradually improved even when masks are not required. Women also reported wearing a mask more often than men. So why are men wearing masks in public places less than women? Perhaps masks are viewed as a sign of fragility or weakness among some men in the USA, as suggested in previous preprint work [22]. Public health messaging that focuses on aligning masks with masculinity would likely be beneficial to improve usage among males in the United States.

It is not surprising that our June data showed that older individuals wear masks more than middle-age and young people because older adults are at higher risk for more severe cases of COVID-19. However, the low percentage of young individuals wearing a mask combined with their potential to be asymptomatic [23] creates problems for case containment. A key and underreported benefit of masks is source control, so mask wearing is important across all age groups. Lower-risk individuals put older adults and those with preexisting conditions of all ages at risk of severe illness by not wearing a mask due to the potential for asymptomatic viral transmission.

One of the most interesting findings of the current study is the evidence of drastically different mask-wearing behavior in rural areas compared with urban and suburban shoppers when mask requirements are not in place. The odds of urban and suburban shoppers wearing a mask was about 4x greater than for rural store-goers during our June data collection period, possibly reflecting the fact that individuals shopping in rural areas perceive lower risk. However, it has been previously reported that mask-wearing habits do not appreciably differ between counties with low and high numbers of COVID-related deaths [14]. Further, while living population density may be lower in rural regions, the size of retail stores is relatively uniform. Thus, differences in the density of shoppers in stores across regions is likely far less than the actual population density differences reflect. Understanding why rural shoppers may resist the use of masks more than shoppers in other areas is an important issue because the health care system in these areas of the United States may be less equipped to handle large spikes in cases. Rural residents may also need to travel further for emergency care, putting them at greater risk for severe consequences of infection. For these reasons, public health messaging that promotes masks in rural communities is very important.

By the last week of July 2020, for stores with no mask mandate, wearing increased to about 80% overall (Fig 4). This may reflect some improvement in voluntary compliance, as many stores had announced mandates to be enforced at later dates. Thus, some individuals may have been moved to comply due to the prospect of needing to wear a mask for future shopping trips. Within these data there were no discernable age-related differences, but it was still evident that more females (82.2%) were wearing masks than males (77.8%). Data after store and the Wisconsin state mask mandates began shows mask wearing increased to over 90% overall (Fig 4).

**Fig 4.**
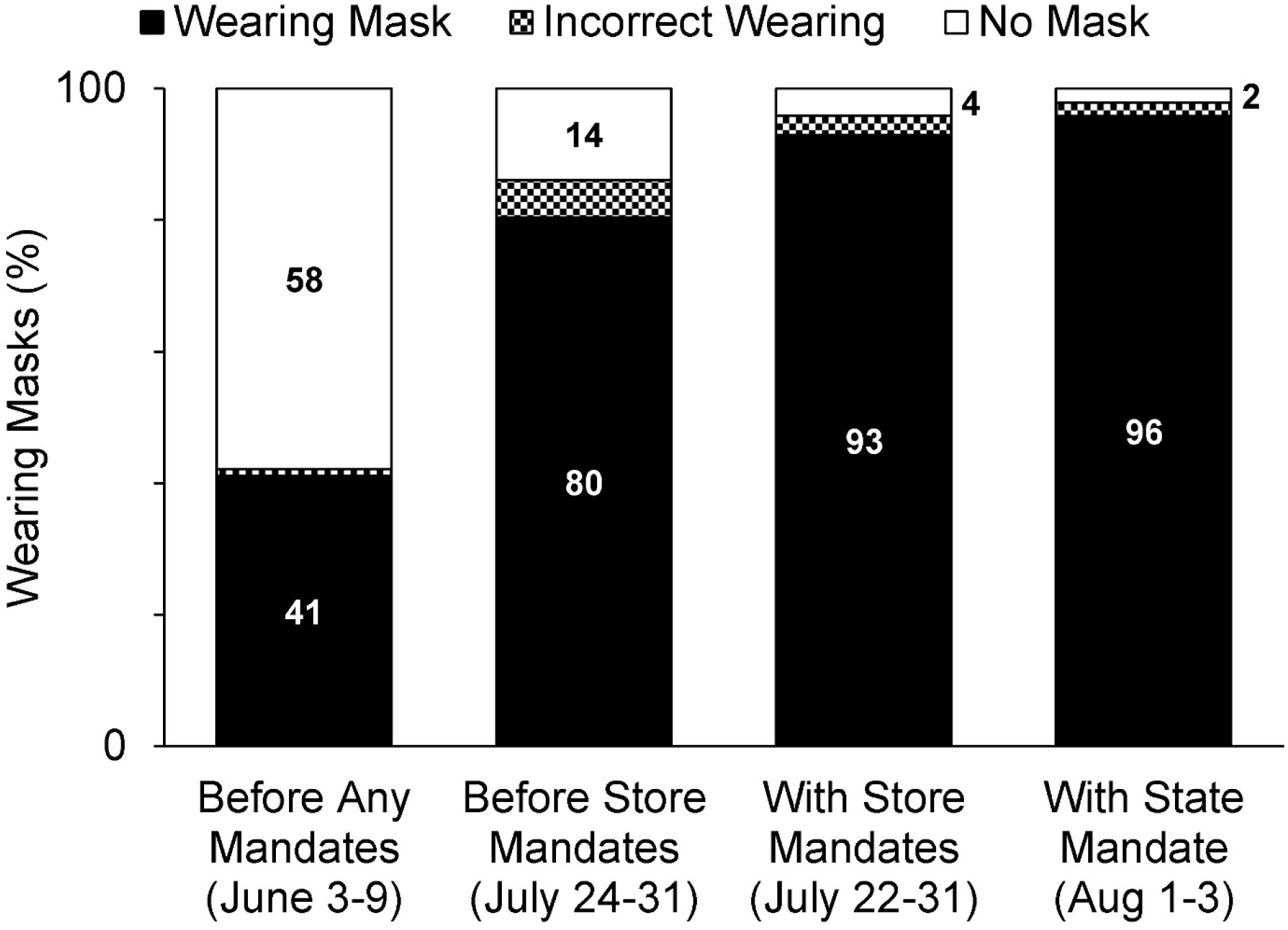
Mask-wearing percentages between June and August 2020. Mask-wearing compliance was poor in June but had improved immediately before mandates began. Compliance rose to > 90% with a mandate in place across all demographics, but a small percentage of the population still resisted. The overlap in “before” and “with store mandates” dates is related to corporate differences in the dates mask mandates began.

Modeling studies suggest mask usage needs to be to nearly universal to have a significant effect on the epidemiological curve [24,25], and our results emphasize that mask mandates are necessary to approach this goal. If public officials do not enact these mandates the welfare of shoppers will continue to be left to retail corporations. While retail store mandates obviously improve mask-wearing compliance among shoppers, our data may indicate that that store regulations could be slightly less effective than mandates enacted by government as a slightly higher percentage of individuals were observed correctly wearing masks after a state mandate began (96% vs 93% with store mandate only).

Mask mandates obviously improve usage, but it deserves mention that a small percentage of the population still resist masks even with retail store and government regulations in place. Though it may be argued that some shoppers might have a health condition that precludes wearing a mask, the Centers for Disease Control and Prevention only recommend against mask wearing if an individual has trouble breathing or is unable to remove a face covering without assistance [26] so it is improbable that this is a valid explanation for all of the mask resistors observed with a state mandate (2%; Fig 4). Mask-wearing mandates are necessary to reduce person-to-person transmission of COVID-19 and save lives as a portion of the general public will resist wearing masks in retail stores without them.

Even with mandates in place, some individuals still resist or wear them incorrectly, putting employees, other shoppers, and themselves at risk due to the potential for viral transmission from asymptomatic carriers [4]. Encouraging voluntary compliance to mask wearing is critical so that retail employees do not have to play the role of “mask police”, risking verbal or physical abuse from resistant shoppers. Close encounters (within six feet) are a great risk to employees if the resistor without a mask is infected. Though some stores provide masks to shoppers after entering, there is still a short period that these individuals are inside of the store without masks on. As the possibility of aerosolized transmission of COVID-19 was recently confirmed [27], every breath and word spoken without a mask on (or without one covering mouth and nose) and especially indoors, increases risk of aerosolized virus spread. Thus, it is critical to wear a mask from entrance to exit of any public indoor space. To further reduce the number of individuals entering without a mask, stores may benefit from providing masks to shoppers that need them outside of the entrance, where possible. Averting cases of COVID-19 will save lives so wearing a mask is a public health service and should not a symbol of frailty or fragility of the individual. Wearing a mask is not about stopping transmission of all cases but minimizing case rates and lessening the public health burden. Importantly, the long-term effects of COVID-19 are still unknown so mask wearing is also vital to reduce burden from future comorbidities that may result within individuals who were infected.

## Conclusions

Masks are an essential public health intervention for combatting the pandemic, and only 41% of individuals observed in our June sample were wearing a mask when entering a retail store. This is less than half the percentage needed to have a significant effect on the reducing the spread of COVID-19 [24,25]. Males, younger individuals, and shoppers in rural communities were observed wearing masks less than other groups. However, with store mandates and a statewide mandate to wear a mask, we showed that mask-wearing compliance rose to over 90% in all groups, including those that resisted mask wearing in June. As officials around the United States continue to debate mask mandates at local and state levels, our data indicated that mandates are necessary to ensure mask-wearing compliance among the public meets the minimum threshold to take control of the COVID-19 pandemic. Our data also highlights that a portion of the general population does not follow public health recommendations without them. Furthermore, even with mandates in place a portion of shoppers (~4%) still resist or wear masks ineffectively. Individual shoppers may consider this information to evaluate the potential risks of in-person shopping based on store and local mask policies, location, and the age and gender of the store’s clientele. Our findings are also important to inform policy makers that mask mandates are necessary to improve compliance to a level that can help flatten the epidemiological curve, saving lives and decreasing burden on the United States health care system.

## Data Availability

Data is available on request.

